# Sex-Specific X-Chromosome Gene Expression in Intracranial Aneurysms

**DOI:** 10.64898/2025.12.08.25341862

**Authors:** Serena Fan, Kerry E. Poppenberg, Vincent M. Tutino, Elad Levy, Daniel Woo, Pui Man Rosalind Lai

**Affiliations:** Department of Neurosurgery, Jacobs School of Medicine and Biomedical Sciences, University at Buffalo, Buffalo, New York, USA; Department of Neurosurgery, Gates Vascular Institute at Kaleida Health, Buffalo, New York, USA; Canon Stroke and Vascular Research Center, University at Buffalo, Buffalo, New York, USA; Jacobs Institute, Buffalo, New York, USA; Department of Radiology, Jacobs School of Medicine and Biomedical Sciences, University at Buffalo, Buffalo, New York, USA; Department of Neurology, Jacobs School of Medicine and Biomedical Sciences, University at Buffalo, Buffalo, New York, USA

## Abstract

**Background:** Women have a higher prevalence of intracranial aneurysms (IAs) than men, but the biological basis for this difference is unclear. Some X-chromosome genes escape inactivation and may contribute to sex-specific susceptibility. This study aimed to identify gene expression differences associated with unruptured IA and to determine whether X-linked gene expressions differ by sex.

**Methods:** Whole-blood RNA sequencing was performed in 166 individuals, including 129 patients with unruptured IAs (108 women and 21 men) and 37 controls (14 women and 23 men). Differential gene expression was evaluated using edgeR with a false discovery rate threshold q-value=*0.05*. Sex-stratified analyses were conducted to study X-linked expression patterns in women and men separately. *CDK16*, the leading X-linked candidate, was validated by real-time quantitative PCR (RT-qPCR) in an independent cohort of 55 individuals with 32 IA cases (20 women and 12 men) and 23 controls (11 women and 12 men).

**Results:** Seven genes were differentially expressed in IA versus controls (*HBG2, AP3B2, COL7A1, CDK16, NUP210L, KDM5C, HSPA1L*), including two X-linked genes: *CDK16* (log_2_FC 0.23, q-value=0.018), and *KDM5C* (log_2_FC 0.32, q-value=*0.04*). *CDK16* showed a strong sex-specific pattern where the expression was higher in women with IA compared to female controls, and no difference was observed between male IA and controls. RT-qPCR confirmed the sex-dependent differential expression of *CDK16* (p<*0.01*).

**Conclusions:** Unruptured IA is associated with distinct blood transcriptional changes, including differential expression of X-linked genes. Additionally, *CDK16* emerges as a potential female-specific biomarker, suggesting a mechanism involving escape from X-inactivation. These findings support further investigation of X-chromosomal regulation in the female predominance of IA and may inform future diagnostic and therapeutic strategies.

## Introduction

Intracranial aneurysms (IAs) are abnormal dilations in blood vessels which can cause devastating hemorrhage if they rupture. Interestingly, women have a higher rate of developing IAs compared to men^1,2^. Both *in vivo* and *in vitro* studies have demonstrated estrogen do not promote IA formation, and in fact, exert a protective effect. Therefore, female hormones do not explain why there is a higher IA prevalence in women^3–5^.

Emerging evidence suggests chromosomal contributions play a role in various neurological, vascular, and immunologic diseases^6–8^. Studies have found that differential gene expression on the X chromosome is important for conferring sex differences in protein expression in several complex disease states including Parkinson’s disease, Alzheimer’s disease, multiple sclerosis, and abdominal aortic aneurysms^6,9–11^. The X chromosome encodes over 800 protein-coding genes and harbors the largest number of immune-related genes in the human genome^12^. While X-chromosome inactivation (XCI) in women (XX) normally balances the dosage of X-linked genes between sexes, approximately 15-30% of X-linked genes escape XCI ^13–15^. This results in differences in gene dosage of X chromosome between the sexes, which is proposed to contribute to sexual dimorphism and women’s enhanced immune susceptibility to immunologic diseases^12,16,17^. More recently, X chromosomal gene dosage differences have also been implicated in ischemic stroke, but this mechanism has not been reported in hemorrhagic stroke or in IA^18^.

In this study, we hypothesize that X-linked genes escaping inactivation in women result in differences in X-linked gene dosage, promoting immune susceptibility and resulting in IA pathophysiology. If so, we expect differential gene expression on the X chromosome comparing patients with IA and without IA. To investigate this, we performed transcriptomic analysis using RNA-seq of blood samples from patients with IAs and controls, and validated an X chromosomal gene, *CDK16*, with RT-qPCR.

## Methods

### Study Population

This study was approved by the University at Buffalo Institutional Review Board (IRB 030-474433), and all participants provided written informed consent prior to sample collection. Patients undergoing cerebral digital subtraction angiography (DSA) at Gates Vascular Institute (Buffalo, NY) between January 2022 and December 2023 were eligible. The study included individuals with unruptured IA identified on noninvasive imaging or follow-up noninvasive imaging of previously detected IAs (IA group), and patients without vascular disease serving as controls (control group). Unruptured IA diagnosis was confirmed by DSA. Exclusion criteria included ruptured IA, prior endovascular or surgical IA treatment, pregnancy, fever >37.8°C, recent surgery, active chemotherapy treatments, autoimmune disease, or immunomodulating therapy. Demographic and clinical data were extracted from patient electronic medical records.

### Blood Collection and RNA Extraction

A 2.5 mL blood sample was collected from the femoral access sheath and transferred into a PAXgene blood RNA tube (PreAnalytiX, Hombrechtikon, Switzerland). Total RNA extraction was performed using the CMG-1084 Chemagic total RNA kit (Revvity, Baesweiler, Germany) according to the manufacturer’s instructions through automated processing on the Chemagic 360 system (Revvity). RNA samples were then run on the Lunatic next-gen UV/Vis spectrophotometer (Unchained Labs) and 4200 TapeStation System (Agilent). RNA integrity was confirmed on an Agilent Fragment Analyzer with the HS RNA assay kit and quantified using the Qubit 4.0 RNA assay (Invitrogen).

### RNA Library Preparation and Sequencing

RNA libraries were prepared using the Illumina RiboZero Total Stranded RNA library prep kit with ribosomal RNA depletion. Total RNA (25-300 ng) was hybridized with rRNA-targeted probes and enzymatically fragmented to 100-200bp. First and second strand cDNA were constructed, followed by adapter ligation, PCR amplification with unique barcodes, and clean-up with AMPure XP beads (Beckman Coulter). Libraries were assessed using the Agilent Fragment Analyzer and Qubit 4.0 High Sensitivity DNA assay. Libraries were pooled to 2nM, denatured, spiked with 1% PhiX control, and sequenced on an Illumina NovaSeq 6000 S2 flow cell (pair-end, 100 bp). Per-cycle basecall (BCL) files generated by the Illumina NovaSeq were converted to per-read FASTQ files using bcl2fastq version 2.20.0.422 using default parameters. The quality of the sequencing was reviewed using FastQC version 0.11.5. Detection of potential contamination was done using FastQ Screen version 0.11.1. FastQC and FastQ Screen quality reports were summarized using MultiQC version 1.5.

### RNA-Seq Analysis

Differential gene expression was analyzed using the edgeR package (Bioconductor v4.7.2) in R. Protein-coding genes present in at least 50% of samples were included. Outliers were identified and removed based on principal component analysis (PCA). Comparisons included IA vs control across all samples, sex-stratified IA (female IA vs male IA), and sex-stratified controls. Subgroup analyses for women and men were also performed. Multiple hypothesis testing correction was performed using the Benjamini-Hochberg false discovery rate (FDR) correction with *q-value*< *0.05*.

### RT-qPCR

Total RNA was reverse-transcribed using a cDNA synthesis kit (ThermoFisher Scientific). The PowerUp™ SYBR™ Green Master Mix (ThermoFisher Scientific) was used for qPCR reactions. The GAPDH primers were from ThermoFisher. The sequence for the GAPDH forward sequence was 5’ ACCACAGTCCATGCCATCAC 3’ and the GAPDH reverse sequence was 5’ TCCACCACCCTGTTGCTGTA 3’. The melting temperatures were 60.20℃ and 59.36℃ respectively. The CDK16 primers were designed using Primer BLAST (NCBI, Bethesda, MD). The forward primer sequence was 5’ GGGACACGTCACGACTATGAA 3’ and reverse sequence was 5’ CCTGCCTCCACCCTCTTATC 3’. The melting temperatures are 59.80 and 59.24 respectively and create a product of 171 base pairs. For validation by RT-qPCR, statistical significance was assessed using Student’s t-test, with *p-values* <*0.05* considered significant.

## Results

### Subject demographics

The RNA-seq discovery cohort included 166 individuals: 129 patients with unruptured IA (108 women, 21 men) and 37 controls (14 women, 23 men). There was no difference in age between groups. Women were overrepresented in the IA group compared to controls (84% vs 38%, p<0.001). Comorbidities including hypertension, coronary artery disease, and diabetes were similar between groups (*p=0.1*, *p=0.23*, *p=0.42*, respectively). Ischemic stroke (8.5% vs 30%, *p=0.002*) and hypercholesterolemia (43% vs 65%, *p=0.042*) were more common in control patients. A family history of IA was more prevalent in IA group (*p=0.006*), while family history of stroke did not differ (*p=0.75*) (Table 1).

**Table 1.** Demographics and clinical characteristics of patients with intracranial aneurysm (IA) and controls in the RNA-seq cohort.

|  | Patients with IA<br>(N=129) | Control (N=37) | Total (166) | *P value* |
| --- | --- | --- | --- | --- |
| Age | 61+14.5 | 65+12.5 | 62+14.1 | *0.07* |
| Sex (Female) (%) | 108 (84) | 14 (38) | 129 (77) | **<0.001** |
| Hypertension (%) | 74 (57) | 28 (76) | 102 (61) | *0.1* |
| CAD (%) | 36 (30) | 15 (41) | 51 (31) | *0.23* |
| Stroke (%) | 11 (8.5) | 11 (30) | 22 (13) | **0.002** |
| High Cholesterol (%) | 55 (43) | 24 (65) | 79 (48) | **0.042** |
| Diabetes (%) | 28 (22) | 11 (30) | 39 (23) | *0.42* |
| Family History of IA (%) | 35 (27) | 2 (5.4) | 37 (22) | **0.006** |
| Family History of Stroke (%) | 19 (15) | 6 (16) | 25 (15) | *0.75* |

### Differential expression in aneurysm patients

RNA-seq analysis identified seven genes significantly differentially expressed between IA patients and controls (*q-value<0.05*): hemoglobin subunit gamma 2, *HBG2* (log_2_FC –2.5, *q-value=1.07×10^−4^*), adaptor related protein complex 3 subunit beta 2, *AP3B2* (log_2_FC –1.79, *q-value*=4.16×10^−4^), collagen type VII alpha 1 chain, *COL7A1* (log_2_FC 2.03, *q-value=0.018*), cyclin dependent kinase 16, *CDK16* (log_2_FC 0.23, *q-value=0.018*), nucleoporin 210 like, *NUP210L* (log_2_FC 1.32, *q-value=0.04*), lysine demethylase 5C, *KDM5C* (log_2_FC 0.32, *q-value=0.04*), and heat shock protein family A, *HSPA1L* (log_2_FC 1.45, *q-value=0.047*) (Table 2). Two X-linked genes, *KDM5C* and *CDK16*, were among these. No differentially expressed genes were found on the Y chromosome. Autosomal genes implicated in vessel trafficking, cytoskeletal organization and extracellular matrix remodeling (*AP3B2*, *COL7A1*) were differentially expressed.

**Table 2.** Seven differentially expressed genes in blood of patients with unruptured intracranial aneurysms (IAs).

| Gene ID | Gene Name | Chromosome Location | log<sub>2</sub>FC | FDR | Up/Down Regulated | Function |
| --- | --- | --- | --- | --- | --- | --- |
| *HBG2* | Hemoglobin Subunit Gamma 2 | 11p15.4 | -2.51 | ***0.1x10<sup>-4</sup>*** | Down | Component of hemoglobin; involved in oxygen transport. |
| *AP3B2* | Adaptor Related Protein Complex 3 Beta 2 Subunit | 15q25.2 | -1.79 | ***0.4 x10<sup>-3</sup>*** | Down | Involved in synaptic vesicle biogenesis |
| *COL7A1* | Collagen Type VII Alpha 1 Chain | 3p21.31 | 2.03 | ***1.8 x10<sup>-2</sup>*** | Up | Structural protein critical for anchoring fibrils in the basement membrane. |
| *CDK16\** | Cyclin Dependent Kinase 16 | Xq26.2 | 0.23 | ***1.8 x10<sup>-2</sup>*** | Up | Serine/threonine kinase regulating vesicle trafficking |
| *NUP210L* | Nucleoporin 210 Like | 1q42.13 | 1.32 | ***0.04*** | Up | Component of the nuclear pore complex |
| *KDM5C\** | Lysine Demethylase 5C | Xp11.22 | 0.32 | ***0.04*** | Up | Histone demethylase involved in transcriptional regulation |
| *HSPA1L* | Heat Shock Protein Family A (Hsp70) Member 1 Like | 6p21.33 | 1.45 | ***0.047*** | Up | Molecular chaperone involved in protein folding and stress response |

\**CDK16* and *KDM5C* are on the X chromosome

To account for sex differences, additional analysis was performed stratified by sex. In the male-only cohort, one gene was differentially expressed (*RAB26*). In the female cohort, three genes were found to be differentially expressed (*AP3B2, CMBL, GAD1*). In women, *CDK16* showed a trend towards increased expression (log_2_FC 0.13, *q-value=0.07*) but this finding was not significant after multiple testing adjusted across all chromosomes (*q-value=0.81*). In male patients, *CDK16* expression was not significantly different (log_2_FC 0.06, *q-value=0.84*). Volcano plots illustrate the distribution of differential expression across the entire cohort and by sex (Figure 1).

**Figure 1.**
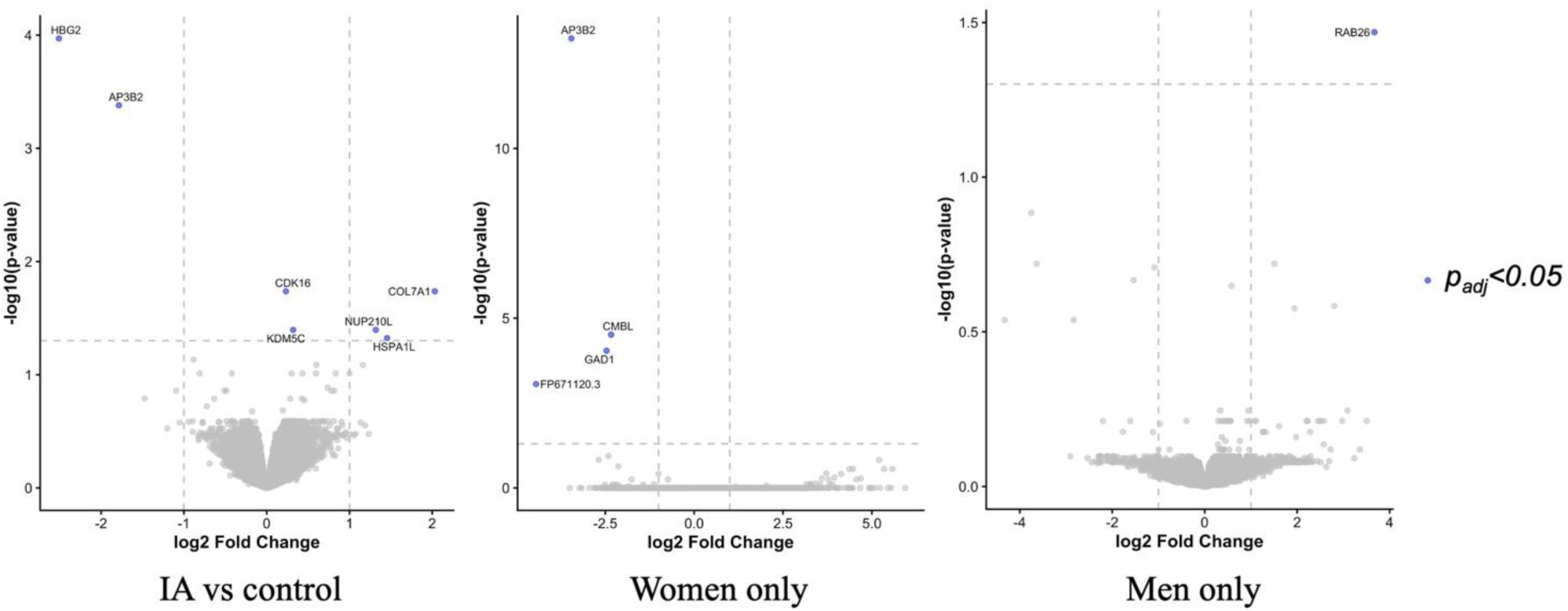
Volcano plot showing differential gene expression (blue) with *FDR-adjusted p-value <0.05* across the entire intracranial aneurysm (IA) vs control cohort, as well as stratified analyses for female-only and male-only cohorts.

### Validation of CDK16 Expression by RT-qPCR

*CDK16*, the top X-linked candidate, was validated in an independent cohort of 55 individuals (32 IA [20 women, 12 men] and 23 controls [11 women, 12 men]). Demographics and comorbidities were similar between groups except for family history of IA (*p=0.048*) (Supplementary Table 1). Consistent with RNA-seq results, *CDK16* expression was elevated in IA patients compared with controls (Gene expression ratio 2.3±0.4 vs 1.2±0.1, *p=0.027*).

Sex-stratified qPCR analysis demonstrated that this difference was driven by females (3.1±0.1 vs 1.1±1.0, *p=0.0095*), whereas expression in males did not differ significantly between IA and controls (1.1±0.4 vs 0.7±0.1, p=*0.37*). Comparing sexes within the IA group, *CDK16* levels were higher in female IA patients than in male IA patients (1.3±0.1 vs 0.3±0.2, *p=0.0002*), while no sex difference was observed in controls (1.6±0.4 vs 0.8±0.4, *p=0.13*) (Figure 2).

**Figure 2.**
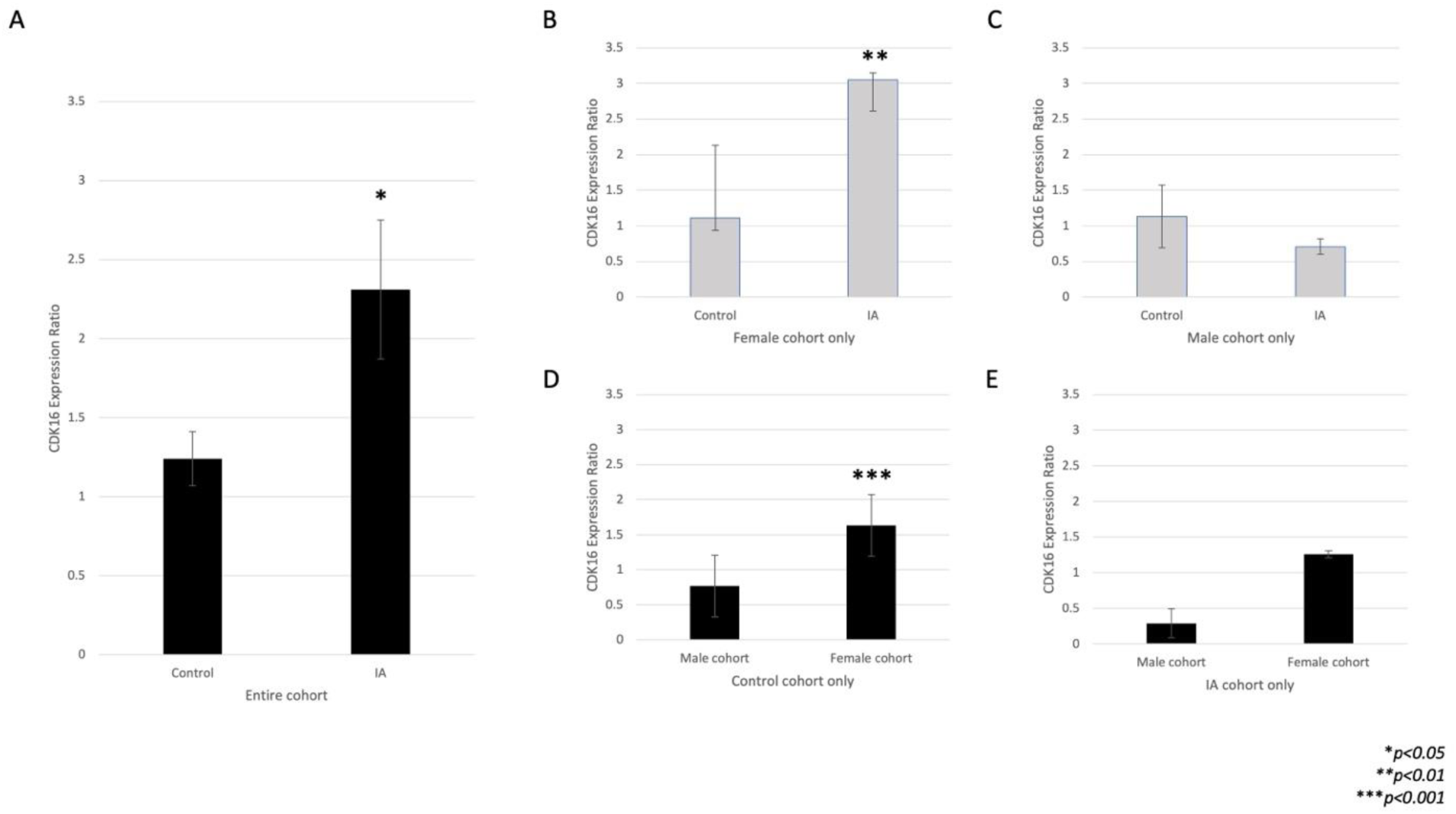
RT-qPCR validation of *CDK16* in intracranial aneurysm (IA) vs control blood samples. *CDK16* expression was significantly elevated in IA compared with control, driven by the female cohort, but no difference was observed in males.

## Discussion

In this study, we identified sex-specific transcriptional differences in patients with unruptured IAs, with a particular focus on X-linked genes. Across the entire cohort, seven genes were differentially expressed between IA patients and controls, including two X-linked genes, *CDK16* and *KDM5C*. Autosomal genes were enriched for pathways related to cytoskeletal organization, vesicle trafficking, and extracellular matrix remodeling, which are consistent with known mechanisms of IA formation including endothelial dysfunction, vascular remodeling and inflammatory signaling.

In women, one X chromosome in each cell is randomly inactivated, and each X-linked gene is expressed from only one allele per cell, creating a mosaic pattern. In contrast, males have only one X chromosome, so all X-linked genes are expressed from one copy. However, up to 30% of X-linked genes may escape inactivation in women, which provide a potential mechanism for heightened inflammatory responses. This may explain stronger immune responses and higher susceptibility to autoimmune diseases in women ^12,17,19^. We hypothesize this heightened inflammatory response, promoted by differential X chromosomal genes in women, explains the female higher risk of IA.

Sex-stratified analyses highlighted that *CDK16* expression differences were primarily driven by female patients. *CDK16*, a member of the Cdc2/Cdkx family of serine/threonine kinases, plays a role in cell cycle regulation and inflammatory signaling through NF-kappa B and cytokine recruitment^20,21^. *CDK16* is known to escape X-inactivation^22^ and is implicated in female-specific immune functions, including the expression of a chimeric RNA, which has been shown to modulate TNF-alpha signaling, further supporting a role in inflammatory regulation^23^. Our qPCR validation confirmed elevated *CDK16* expression in women with IA, whereas expression in men did not differ significantly, supporting a sex-dependent effect. To our knowledge, this is the first evidence of differential X-chromosome gene expression in IA, suggesting a novel molecular pathway that may contribute to the female predominance of IA formation.

In addition to X-linked genes, we found differential expression of autosomal genes involved in cytoskeletal organization, vesicle trafficking, and extracellular matrix remodeling, including *AP3B2* and *COL7A1*. These pathways are consistent with known mechanisms of IA formation, including endothelial dysfunction, vascular remodeling, and inflammatory signaling^24–27^. Chronic inflammation, triggered by factors such as hypertension, and atherosclerosis, weakens endothelial wall, thereby promoting IA formation^28,29^. Sex hormones, such as estrogens, protect against endothelial damage and suppresses pro-inflammatory cytokines such as *IL-6* and *TNF-alpha*, while increasing anti-inflammatory *IL-10*^30,31^. Post-menopausal estrogen has been known to be associated with increased proinflammatory cytokines, such as *IL-1*, *IL-6*, and *TNF-alpha*, thereby exacerbating inflammatory responses in women^32,33^. This may explain why hormonal effects alone cannot explain the observed sex differences in IA, but the effect becomes more apparent after the loss of estrogens.

There are growing literature suggesting that neurologic and vascular diseases are associated with the X chromosome ^6–8^. Our findings support this broader concept that the X chromosome contributes to sex differences in vascular and inflammatory diseases. Prior studies have demonstrated sexually dimorphic expression of X-linked genes, including *KDM5C* and *KDM6A*, in stroke and other cardiovascular conditions^34^. Our study also found *KDM5C* to be differentially expressed in IA formation. X-chromosome genes, such as *KDM5C* and *CDK16*, may act in concert with hormonal and inflammatory mechanisms to modulate vascular susceptibility, predisposing women to IA formation.

Our study has several limitations. It is observational and cannot establish causality between gene expression and IA pathophysiology. The sample size, particularly in the sex-stratified analyses, is relatively small, which may limit the detection of subtle expression changes. RNA profiling was performed in blood and may not fully reflect transcriptional changes within IA tissues. Further studies are needed to confirm the mechanistic role of *CDK16* and other X-linked genes in IA formation. Additionally, sex differences in gene expression does not account for patient comorbidities or environmental factors, such as smoking, hormonal status, which were not controlled in this study. Future work will require larger, longitudinal cohorts to better define mechanistic contributions of these genes and their role in sex-specific effects.

## Conclusion

Our findings demonstrate sex-specific differences in gene expression associated with unruptured IA, particularly involving X-linked genes. *CDK16* emerges as a compelling candidate biomarker in women, consistent with its known escape from X-inactivation and potential modulation of inflammatory pathways. These results provide a foundation for future mechanistic and translational studies in understanding female predominance of IA and in developing sex-specific diagnostic and therapeutic strategies.

## Data Availability

All data supporting the findings of this study are included in the manuscript and supplementary materials. Additional raw data are available from the corresponding author upon reasonable request

## Acknowledgements, Sources of Funding, & Disclosures

## Acknowledgements

Acknowledgements

The authors thank the patients that participated in this study. We also thank the Congress of Neurological Surgeons for supporting this research.

## Abbreviations

*CDK16*: cyclin-dependent kinase 16
IA: intracranial aneurysm
*KDM5C*: lysine demethylase 5C
log_2_FC: log fold change
qPCR: quantitative polymerase chain reaction
q-value: FDR-adjusted p-value
RNA: ribonucleic acid
RNA-seq: RNA sequencing

